# Revising and evaluating the Safe Recovery fall prevention education program with patients and staff in a hospital rehabilitation setting: A mixed methods study

**DOI:** 10.1101/2023.06.23.23291842

**Authors:** Jacqueline Francis-Coad, Melanie K Farlie, Terry Haines, Tammy Weselman, Linda Black, Philippa Cummings, Anne-Marie Hill

## Abstract

**Background:** Providing patients with falls prevention education can improve their overall safety and reduce their risk of falling in hospital. Partnering with patients and staff in developing and evaluating such programs could better enable patient learning and translation of safety messages.

**Aim:** To create a revised version of the Safe Recovery falls prevention education program (SRP) in partnership with patients and hospital allied health staff, to improve patient engagement in undertaking strategies to reduce their risk of falling in hospital.

**Methods:** Two-phase sequential mixed methods participatory design. In phase 1 patient (n=10) and staff (n=10) consumer engagement surveys and discussions were undertaken to inform program revision. New resources (video and workbook) were co-produced and staff were trained to deliver the revised program to patients. In phase 2 patients (n=10) were surveyed pre and post revised program delivery and staff were surveyed regarding their reaction to the revised program. Deductive content analysis and Wilcoxon Signed Rank Tests were used respectively to analyse qualitative and quantitative findings.

**Results:** Patients and staff were very satisfied with the revised program, with patients demonstrating significant improvements in knowledge, awareness, motivation and intention to reduce their risk of falling. Staff perceived that the revised resources showed significant improvements in aesthetic appeal and ability to engage patients in learning.

**Conclusion:** Patients and staff contributed to successfully revising the Safe Recovery program with positive reactions to the co-produced resources. Participating in the revised program significantly improved patients’ knowledge and attitudes to reduce their risk of falling. Investigating the impact of the revised program on patients’ behaviour change and on reducing hospital falls is warranted.

## INTRODUCTION

Falls and injuries from falls remain the most frequently reported safety incidents in hospital inpatient settings. Approximately 30%-50% of falls in hospital result in physical injury including fracture (1-3%) and head injuries, with negative psychological consequences and delayed functional recovery.^1–4^ This can increase patient length of stay and subsequently escalate hospital costs,^4^ therefore a priority for all hospitals is preventing falls and injurious falls.

It has been hypothesised that a discrepancy between a patient’s perceived personal falls risk and their actual level of risk contributes to patient falls in hospital.^5, 6^ This ‘mismatch’ may occur for numerous reasons, such as a patient misperceiving their current level of function and capability, not seeking staff assistance or possessing a false sense of security whilst being in hospital.^5, 6^ Patients may also have limited understanding of how falls occur in hospital and the relevant falls risk factors that they should address, such as using a gait aid to compensate for poor balance.^7–9^ A misperception of personal risk and lack of understanding often results in patients engaging in mobility tasks that they are unable to complete safely during a hospital admission, resulting in a fall.^5, 6, 10^ These types of falls may be preventable if patients are provided with, and adhere to, falls prevention advice.

While multifactorial strategies, such as comprehensive risk assessment, are recommended in hospital settings, world fall guidelines have recently recommended that fall preventive patient education should be provided for older patients to specifically target deficits in patient awareness and comprehension of personal falls risk and provide strategies to help prevent falls in hospital.^11^ A recent systematic review of interventions to reduce falls in hospitals confirmed the importance of patient education. This review identified that evidence-based patient education was the only intervention, from a range of common strategies examined, that was effective at reducing patient fall incidence and fall rates.^12^ Falls prevention education programs that incorporate educational and behavioural change theory, active learning designs, and combinations of education delivery modes have been shown to be more likely to produce successful outcomes.^13^ In addition, new insights for developing education for older adults highlight attending to human information processing, such as minimising cognitive load and chunking related information into clusters, in order to deliver effective learning outcomes.^14^ Therefore, hospital fall prevention education programs for patients should be designed incorporating these components and include evaluation of outcomes.

The Safe Recovery Program (SRP), a hospital patient education program developed in Australia, has demonstrated effectiveness in reducing falls rates by 40% and fall related injuries by 35% in a large randomised controlled trial conducted in hospital rehabilitation units.^15^ Briefly, the SRP provides individualised fall prevention patient education, including active learning (goal setting), delivered by trained allied health professionals. These staff engage in personalised discussion with patients, supported by use of multi-media resources (video and patient workbook).^16^ The program outlines when, where and why falls happen and three simple steps to prevent falls in hospital; knowing if you need help (raising risk awareness), asking for help and waiting for help to arrive.^16^ The SRP was developed across three large randomised controlled trials,^15–17^ with the most recent trial using a clustered ward-level approach to deliver the patient education program alongside training and feedback to staff.^15^ However, the program resources created in 2009 were based on the original trial during which subgroup analysis indicated that there may be benefit for cognitively impaired patients as well as for those with good cognition.^17, 18^ The original SRP was deliberately designed to inclusively cater for people with cognitive impairment and included very slow speech progression and broadly set out simple graphics in the written materials. The first SRP trial indicated that while there was benefit for patients with mild levels of cognitive impairment it was not effective for patients with moderate or severe cognitive impairment.^16^ Therefore the later cluster trial delivered the SRP program on participating wards to all patients with no or mild cognitive impairment.^15^ The ward level delivery of the SRP significantly reduced falls in participating wards compared to control wards. Importantly, patients with cognitive impairment on participating wards who did not directly receive the education benefited from the reduced falls rates. This was most likely because the ward delivery impacted safe culture including through adaptations of the environment and changes in staff practice.^9, 19^

While the SRP was effective, technology for producing and delivering patient education has progressed and continues to evolve, therefore engagement with stakeholders and consumers (hospital staff and patients) regarding their reactions and current preferences is now required.^14^ This is important as the accessibility and quality of an education program can influence patient engagement and subsequent learning outcomes.^20^ Therefore, the aims of this study were:

i. To create a revised version of the resources for the Safe Recovery fall prevention education Program with staff and patients, informed by their reactions to and learnings from the original program and
ii. To evaluate staff and patient reactions to and learnings from the revised version of the SRP at a hospital rehabilitation unit

## METHOD

### Design

A sequential mixed method design^21^ using a participatory approach was conducted in two phases between May 2021 and July 2022 (*see Figure 1 below*). Participatory approaches engender the core philosophy of ‘inclusion’ meaning stakeholders and consumers have valued engagement in the research process in partnership with the research team.^22^

**Figure 1.**
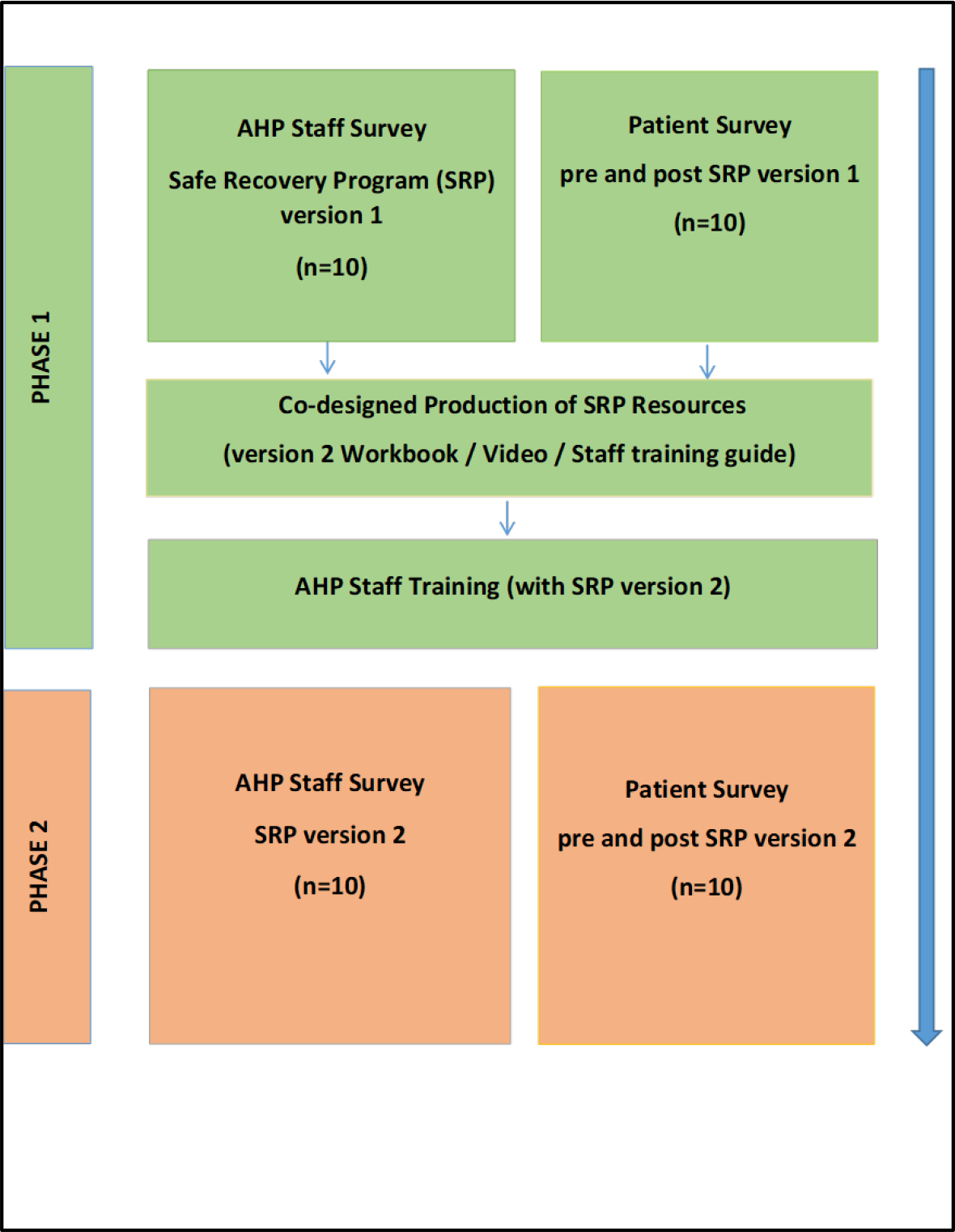
Sequential Mixed Methods Design

Ethical approval was obtained from the Human Research Ethics Committees of Ramsay Health Care WA/SA (2126W) and the University of Western Australia (2021/ET001151). All participants provided written informed consent.

### Setting and participants

The study was conducted at a 31-bed private hospital in metropolitan Perth, Western Australia. The hospital provided inpatient rehabilitation services, including physiotherapy and occupational therapy, for post-surgical and general medical patients following discharge from acute care settings. Patients received the SRP (original version) delivered by trained allied health professional (AHP) staff as part of usual care.

Participants were AHP staff and patients. Eligibility criteria for staff were being employed in an AHP role and the ability to communicate in English with a minimum of three months experience delivering the SRP to patients. Eligibility criteria for patients were being aged 18 years and above, admitted for rehabilitation and ability to read and communicate in English. The AHP manager used the hospital electronic records to obtain a purposive pilot sample that represented the age range, gender and ethnicity of the hospital population.

### Phase 1 procedure

All AHP staff undertaking the role of SRP patient educators, who had consented, completed an online survey seeking their reactions to the SRP. Concurrently, patients meeting the eligibility criteria who consented to participate completed a pre-program survey to ascertain their perceptions and baseline knowledge of fall prevention in hospitals. All patient surveys were conducted face-to-face with a trained research assistant (RA) who recorded responses verbatim on a laptop computer.

AHP staff then delivered the SRP to patients face-to-face within 24 hours of their admission. The original SRP comprised a 15-minute video played on a portable DVD player and paper patient workbook (A4-sized), with large font size, outlining hospital falls epidemiology, fall risk factors and fall prevention strategies. The content was designed to meet grade six reading level as per recommended guidelines for patient education materials.^23^ Patients were asked to review and reflect on the SRP before writing their personal goals in the workbook. AHP staff then returned the next day to discuss goals and proposed strategies to prevent falls whilst in the hospital with the patient. Three to four days after receiving the SRP, the RA returned to conduct the post-program survey to gain patients’ reactions to and learning from the SRP.

### Co-design of the revised SRP

A transcript of findings from the AHP staff and patient surveys was provided electronically to phase 1 participants for comment. A sub-group of three AHP staff and two patients volunteered to participate in a final review of proposed SRP revisions together with the research team prior to production. All stakeholders, including AHP managers and staff, professional videographer, patient actors, and the research team met iteratively, both on-site (during filming) and online during the program revision process.

AHP staff delivering the revised SRP attended a one hour face to face training workshop conducted by the lead researcher. The training included the latest evidence on hospital falls prevention and instruction on how to deliver the revised SRP using the new resources.

### Phase 2 procedure

Following the delivery of the revised SRP participating AHP staff completed an online survey seeking their reactions to the revised program. A second group of patients were recruited as described in phase 1 to evaluate the revised SRP. Those consenting completed the pre-program survey as described in phase 1. AHP staff delivered the revised SRP as described in phase 1 but using the new video played on a tablet and the new patient workbook. Three to four days after receiving the revised SRP, the RA returned to conduct the post-program survey to document patients’ reactions to and learning from the revised program.

### Outcomes measured in Phases 1 and 2

Participants’ (AHP staff and patients) responses to both versions of the SRP were evaluated using surveys that included a mix of closed, open and Likert scale responses. Two key outcomes were measured ‘Reaction’ to the SRP and ‘Learning’ from the SRP. ‘Reaction’ explored to what degree participants reacted favourably to the program. This included patients’ views on the ability of the program to engage them in its content, its relevance, and satisfaction with the program delivery and learning resources. ‘Learning’ explored patients’ development of falls knowledge and perceived level of risk for falls along with levels of awareness, confidence, motivation and commitment to undertake falls prevention strategies.

The theoretical underpinning for the SRP evaluation was based on the first two levels of the New World Kirkpatrick Model for evaluating training programs, which evaluate learners’ reaction and learning to the training.^20^ The Kirkpatrick model has been used as a systematic framework to appraise training in different fields such as simulation-based training^20^ and medical education.^24^ This framework^20^ guided the development of staff and patient questionnaires along with questionnaires published in prior evaluations of the SRP.^15–17^

## Data analysis

### Qualitative data

Qualitative responses from open text items in the patient and AHP staff surveys were read multiple times by the researchers (JFC, TW) for familiarity and managed using NVivo software (QSR International Pty Ltd., Version 12, 2018). Deductive content analysis was selected to evaluate the SRP as there was prior knowledge regarding the research topic, however, a revised version of the program was now being evaluated.^25^ AHP staff and patient responses appraising the original and revised versions of the SRP were mapped to the New World Kirkpatrick Model by constructing a category matrix using the criteria describing ‘Reaction’ to, and ‘Learning’ from the program.^20^ Confirmability of qualitative findings was ensured through the use of verbatim participant quotations.^26^

### Quantitative data

Quantitative survey data were entered into Microsoft Excel (Microsoft Office 2019) and analysed using SPSS version 27 statistical software package (IBM SPSS Inc., Chicago, IL, USA). Descriptive statistics (frequencies/percentages) were used to present participant demographics, frequency of content responses and Likert scale responses (Strongly Disagree, Disagree, Undecided, Agree, Strongly Agree). Differences between AHP staffs’ reactions to the original and revised versions of the SRP were examined using a Wilcoxon signed-rank test.^27^ Differences between phase 1 patient learnings pre and post participation in the original SRP and differences between phase 2 patient learnings pre and post participation in the revised SRP were examined using Wilcoxon signed-rank tests.^27^ Content from all data sources were combined in a content cloud, a type of visualisation tool for comparing and summarising information, to clarify the priority findings.^28^ The study was reported using the Guidance for Reporting Involvement of Patients and the Public (GRIPP) guidelines for reporting patient and public involvement in health and social care research.^29^

## RESULTS

### Participant characteristics

#### AHP Staff

Ten allied health staff undertaking the role of SRP educator participated, eight (80%) being female with a mean age 31.2 years (SD ± 7.8 yrs). Six (60%) were physiotherapists and four (40%) occupational therapists, six (60%) had five or more years professional experience and eight (80%) had been SRP educators for more than one year.

#### Patients

Twenty patients participated as two independent groups of ten, the first group in phase 1 and the second group in phase 2. The key reason for patients’ admissions was for rehabilitation following surgery (n= 17, 85%) with 15 (75%) having experienced one or more falls in the past 6 months. Additional participant characteristics are shown in *Table 1 above*.

**Table 1.** Patient characteristics

| Characteristic | Patient group (n=10)<br>completing original<br>SRP<sup>a</sup> | Patient group (n = 10)<br>completing revised<br>SRP<sup>a</sup> |
| --- | --- | --- |
| Age range (years) | 59 - 88 | 59 – 83 |
| Sex female n (%) | 8 (80) | 3 (30) |
| Ethnicity n (%) |  |  |
| Caucasian | 9 (90) | 9 (90) |
| Asian | 1 (10) | 1 (10) |
| LOS<sup>b</sup> in days, range (mean) | 7 – 19 (11.20) | 6 – 18 (12.10) |
| Condition being rehabilitated n (%) |  |  |
| Non-surgical spinal | 2 (20) | 1 (10) |
| Post spinal surgery | 2 (20) | 2 (20) |
| Post abdominal surgery | 1 (10) | 0 (0) |
| Post total hip replacement | 1 (10) | 2 (20) |
| Post total knee replacement | 3 (30) | 2 (20) |
| Post lower limb fracture | 0 (0) | 3 (30) |
| Post upper limb fracture | 1 (10) | 0 |
| History of falls past 6 months, n(%) |  |  |
| None | 2 (20) | 3 (30) |
| One | 2 (20) | 5 (50) |
| 2 – 4 | 6 (60) | 2 (20) |
| Type of walking aid used n (%) |  |  |
| None | 0 (0) | 1 (10) |
| 2 wheeled walker | 6 (60) | 9 (90) |
| 3 wheeled walker | 1 (10) | 0 (0) |
| 4 wheeled walker | 2 (20) | 0 (0) |
| Gutter frame | 1 (10) | 0 (0) |
| Notes: <sup>a</sup> Safe Recovery Program, <sup>b</sup> Length of stay |  |  |

## Phase 1

### Reactions to SRP original version

#### AHP Staff

Overall staff were somewhat dissatisfied with the original SRP, namely the video and workbook resources, with one (S1) summing them up as “*a bit basic and slightly outdated.*” Primarily, staff attributed low levels of patient engagement with the video resource to the predominantly still photography slide format, low resolution images and poor audio quality. Staff also commented that the video was less engaging for patients with higher levels of cognition and normal hearing as the pace was deemed too slow (S3) “*they don’t feel like it relates to them at their level of function…they can find it quite frustrating to sit through the full-length video.*” Low engagement with the workbook was attributed to the language level used, one (S2) commented “*the language in the video/book needs re-writing as it can seem a bit childish.*”

Staff felt the concept of the SRP educating patients to prevent falls whilst in hospital was relevant, as in their experience many patients had not received any personalised falls prevention education prior to admission, one reflected (S4) “*Some patients find it very educational.”* Another added (S7) that the SRP messages were “*on the mark*” as they “*encourage the patient to pro-actively reduce their risk of falling.*”

#### Patients

Patients (P1-10) expressed low levels of satisfaction with the original SRP resources, particularly the video, one (P6) perceived “*it was childishly done.*” The slow pace and length of the video limited engagement, with one patient (P2) stating “*I think a lot of people would get half-way through this and not bother with the rest.*” Delivery of the video on a small DVD player screen also inhibited engagement as it was challenging for some older people with lower levels of vision to view with ease, one commented (P1) “*it was a small dark video…must be 20 years old or more, no one would be engaged in this.*” Some patients thought the A4 sized workbook was (P4) “*too big*” to handle comfortably and one (P7) pointed out “*there was a lot of white space*” and consequently too many pages. This reduced their desire to engage in reading in full, with one surmising she (P4) “*wasn’t enthused.*” However, program relevance had mixed perceptions. Some patients acknowledged they were at risk of falling as (P9), who reflected “*it did accentuate the problem of falls*” but others did not identify with being ‘at risk’, one added (P2) “*it was informative, but it doesn’t apply to me.*”

### Learnings from SRP original version

#### Patients

Hospital falls epidemiological knowledge showed significant improvement following delivery of the original SRP (p=0.010) along with patients’ awareness of the measures needed to reduce their risk of falling in hospital (p=0.008), as shown in *Tables 2 and 3 respectively below*.

**Table 2.** Comparison of patients’ falls epidemiological knowledge pre and post the Safe Recovery Program (both original and revised versions)

| Patients | Pre-test correct<br>responses<br>Median (IRQ) | Post-test correct<br>responses<br>Median (IRQ) | Wilcoxon signed<br>rank test<br>p value |
| --- | --- | --- | --- |
| Group completing the<br>original SRP (n=10) | 0 (0-1) | 2 (1-2) | 0.010 |
| Group completing the<br>revised SRP (n=10) | 1 (1-1) | 3 (2-3) | 0.006 |
| Note: Knowledge questions<br>i. When (time of day) do falls occur most frequently in hospitals?<br>ii. Where (location) do falls occur most frequently in hospitals?<br>iii. For every 100 patients on this ward, how many do you think would fall before they leave? (Correct answer + or – 5)<br>iv. For every 100 falls on this ward, how many do you think would result in a physical injury? (Correct answer + or – 5)<br>v. For every 100 falls that occur on this ward, how many do you think are witnessed by a hospital staff member? (Correct answer + or – 5) |  |  |  |

**Table 3.** Comparison of patients’ attitudes to falls prevention pre and post participation in the original Safe Recovery Program

| Item | S / D<br>Pre<sup>a</sup> /<br>Post<sup>b</sup> | D<br>Pre<sup>a</sup> /<br>Post<sup>b</sup> | UN<br>Pre<sup>a</sup> /<br>Post<sup>b</sup> | A<br>Pre<sup>a</sup> /<br>Post<sup>b</sup> | S / A<br>Pre<sup>a</sup> /<br>Post<sup>b</sup> | p value |
| --- | --- | --- | --- | --- | --- | --- |
| For me, taking measures to reduce my risk of falling would be useful in hospital | 0 / 0 | 0 / 0 | 0 / 0 | 5 / 2 | 5 / 8 | 0.083 |
| Most people whose opinion I value approve of me taking measures to reduce my risk of falling in hospital | 0 / 0 | 0 / 0 | 0 / 0 | 5 / 4 | 5 / 6 | 0.705 |
| I feel positive about reducing my overall risk of falling in hospital | 0 / 0 | 0 / 0 | 1 / 0 | 6 / 2 | 3 / 8 | 0.058 |
| I am aware of the measures needed to reduce my risk of falling in hospital | 0 / 0 | 0 / 0 | 0 / 0 | 9 / 2 | 1 / 8 | **0.008** |
| I am confident that if I wanted to, I could reduce my risk of falling in hospital | 0 / 0 | 0 / 0 | 2 / 0 | 6 / 5 | 2 / 5 | 0.059 |
| While I'm in hospital, I intend to undertake measures to reduce falls or my risk of falling | 0 / 0 | 0 / 0 | 0 / 1 | 4 / 4 | 6 / 5 | 0.414 |
| I have a clear plan of how I will take measures to reduce falls or my risk of falling in hospital | 0 / 0 | 0 / 0 | 1 / 1 | 6 / 4 | 3 / 5 | 0.527 |
| Notes: <sup>a</sup> Pre- original Safe Recovery Program / <sup>b</sup> Post- original Safe Recovery Program<br>SD= strongly disagree, D=Disagree, UN= undecided, A=agree, SA= strongly agree |  |  |  |  |  |  |

### Changes to the SRP

Engagement of a professional videographer enabled better quality of captured images, lighting, audio integration and execution to enhance delivery of the program messages. The new narrative style using a content presenter in a real and operational hospital setting provided a more authentic and contemporary context. The video was shortened to eight minutes and delivered using a single click on a tablet device. The workbook was reduced to an A5-sized booklet format, text phrases were shortened and bolded in colour, reducing repetition and white space. New contemporary photographic images were utilised that mirrored the text and video messaging.

## Phase 2

### Reactions to SRP revised version

#### AHP Staff

Staff perceived the new resources were contemporary, engaging and easy for patients to understand. Staff reaction ratings to the revised SRP showed significant improvements in the program’s ability to utilise multimedia effectively (p=0.007), easily access the video (p=0.007) and workbook (p=0.041), provide instructions and demonstrations that reflected clinical practice (p=0.007) and explain how to prevent falls in hospital (p=0.011) as shown in *Table 4 below*.

**Table 4.** Comparison of AHP staff reactions to the original and revised versions of the Safe Recovery Program

| Item | S / D<br>Ov<sup>a</sup> /<br>Rv<sup>b</sup> | D<br>Ov<sup>a</sup> /<br>Rv<sup>b</sup> | UN<br>Ov<sup>a</sup> /<br>Rv<sup>b</sup> | A<br>Ov<sup>a</sup> /<br>Rv<sup>b</sup> | S / A<br>Ov<sup>a</sup> /<br>Rv<sup>b</sup> | p value |
| --- | --- | --- | --- | --- | --- | --- |
| The SRP used an appropriate range of media to illustrate the key concepts | 0 / 0 | 5 / 0 | 2 / 0 | 3 / 5 | 0 / 5 | **0.007** |
| The video learning was presented in an easily accessible format | 1 / 0 | 4 / 0 | 2 / 0 | 3 / 5 | 0 / 5 | **0.007** |
| The workbook learning was presented in an easily accessible format | 0 / 0 | 3 / 0 | 2 / 0 | 5 / 9 | 0 / 1 | **0.041** |
| The instruction and demonstration in the SRP reflect the way that you would apply falls prevention strategies in practice | 0 / 0 | 3 / 0 | 5 / 0 | 2 / 5 | 0 / 5 | **0.007** |
| Overall, the SRP adequately explains to patients how to prevent falls in hospital | 0 / 0 | 3 / 0 | 4 / 0 | 2 / 3 | 1 / 7 | **0.011** |
| Notes: <sup>a</sup> Original version of Safe Recovery Program / <sup>b</sup> Revised version of Safe Recovery Program<br>SD= strongly disagree, D=Disagree, UN= undecided, A=agree, SA= strongly agree |  |  |  |  |  |  |

Improved patient engagement with SRP resources was attributed to (S6) “*clearer messaging*” delivery with “*more modern content and vibrant audio*” making it more enjoyable for patients. Staff felt the new video had a better narrative structure, one stated (S3) “*the presenter talks slower but still at a speed that doesn’t make viewers feel demeaned or that their own knowledge and experience were underestimated”* another added (S4) “*its timely and not too long, avoiding inattention and distraction.*” The updated workbook was deemed an improvement on the original due to being smaller and less bulky one commented (S2) “*it’s a good size to have on a tray table.*” The graphics were thought to be sharper with good use of supporting text in bold and upper case, with one staff member (S7) validating, “*it better emphasised points* [SRP messages].” Program relevance was heightened, one staff member commented (S10) “*it’s absolutely appropriate to our patient population as the majority have had falls in the past or are at high risk because they are below their baseline mobility.*” Another added (S3) “*clear instruction of when they are most at risk is valued information for any vulnerable patient.*” Staff perceived patients to feel safer after participating in the revised SRP as they were, as one reported (S8) “*more aware of their risks and the strategies they can employ to reduce falls.*”

#### Patients

Patients (P11-20) expressed high levels of satisfaction and enjoyment following participation in the revised SRP, one (P13) reflected “*It all made sense, it was easy to follow…there was a story there.*” The program made patients feel safer and SRP messages were deemed (P15) “*sensible…and well reinforced using different contexts*” giving them broad appeal. The video received very positive feedback from patients as they felt it was ‘well put together.’ They found the video interesting and easy to watch, the pace was good, and it kept their attention throughout, one patient concluded (P18) *“…nice and clear and not too long, otherwise people tend to lose interest.*” Patients appreciated the explicit language used as it was in layman’s terms and not overly complicated. Overall, the video received the highest praise with patients loving the narrative style presentation one (P16) summed up “*the presenter did a great job*…*it was a damn good video!*”

Responses to the new patient workbook were enthusiastic and positive, patients believed it to be a valuable resource for anyone in a hospital setting, especially for those who preferred reading something. Patients praised the workbook for its manageable size, one explained (P13) “*it’s a nice size to hold in your hands*” with clear easy to read print and the use of contrasting bright colours that made the information stand out, one patient remarked (P12) “*it had good readable font size and shaded text boxes*” another affirmed (P17) “*I can read it easily after my cataract surgery..* The pictures in the book were perceived as helpful in reinforcing the safety messages as they illustrated the text, one patient stated (P14) “*the use of graphics and pictures adds interest…it reinforces what you should do.*” Reinforcement in written format was perceived beneficial, one patient added (P11) “*Older patients like me may need reinforcement…as my generation are not used to treating a screen as a medium for learning*”. Patients also reported that the information was presented in a logical order and points were repeated for emphasis, making it a good reminder for them. They also liked the concept of setting their goals and writing them down, as it got them involved in the process of fall prevention.

The SRP was deemed relevant for everyone in a hospital setting. Most patients identified being at risk of falling whilst recovering in hospital, one pointed out (P14) “*it was highly relevant because you don’t feel like your usual self*” another reflected (P20) “*I could relate to several of those messages…my legs are weak, and I’ve been light-headed and dizzy.*” The program also provided reassurance for patients during recovery, with one summing up (P13) “*It makes you feel you’re not alone in your predicament.*”

### Learnings from SRP revised version

#### Patients

Patients receiving the revised SRP showed significant improvements in their falls knowledge (p=0.006) (*see Table 2 above*). Patients’ perception (p=0.046), awareness (p=0.005), motivation (p=0.020) and intention (p=0.024) to undertake strategies to reduce their risk of falling in hospital was also significantly improved *(see Table 5 below*).

**Table 5.**
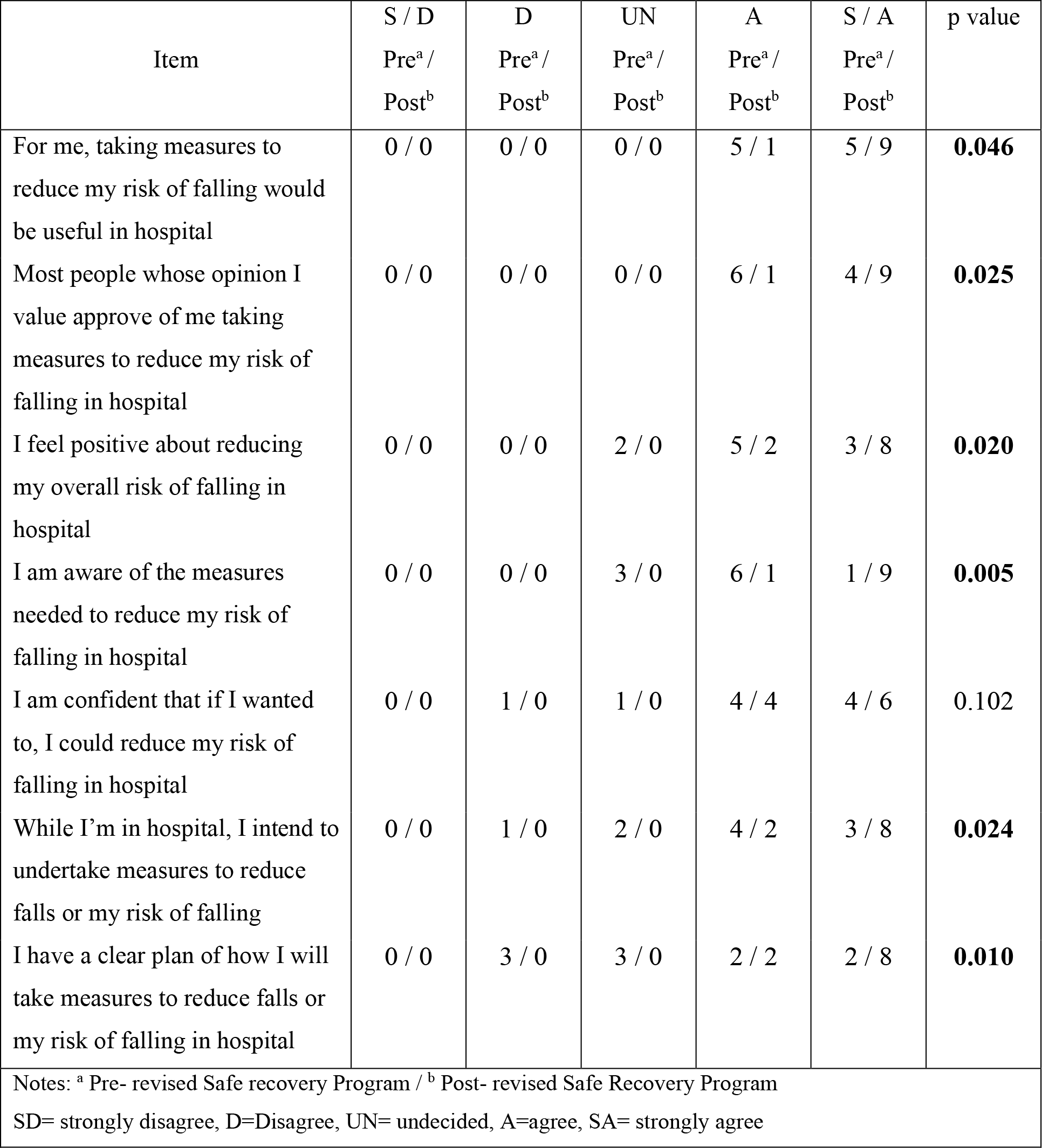
Comparison of patients’ attitudes to falls prevention pre and post participation in the revised Safe Recovery Program

The revised SRP provided patients with a clearer plan (p=0.010) about how to reduce their risk of falls in hospital. Male patients learned it was acceptable to ask for help, one disclosed (P19) “*Guys shouldn’t be proud to ask for help*” another affirmed (P16) “*Don’t be a hero! particularly us blokes, press the bloody bell and be patient* [laughter].” Patients demonstrated improved risk and strategy awareness one stating (P20) “*I didn’t know…that if you feel dizzy you should tighten your leg muscles!”* another concluded (P15) “*despite what I might think I am at significant risk of a fall…but three things, know if I need help, ask for help and wait for help can ensure I’m safe!*”

## DISCUSSION

This research resulted in a revised version of the SRP successfully co-designed by patients, staff and researchers. The updated resources were acceptable to patients and staff, particularly the new contemporary video resource. Patients’ reactions and learning to both the video and the workbook demonstrated better engagement with the SRP messages whilst recovering in hospital.

Staff and patients as program consumers were able to highlight the program areas in need of improvement and contributed to the successful co-design. Patient and staff feedback emphasised that it was particularly important to update the video resource. This feedback was supported by the researchers, based on the evidence from the original SRP about the importance of the video for patient learning about hospital falls. The original SRP resources were piloted in a RCT that evaluated patients’ reactions and learnings about falls when provided with the video presentation compared to being provided with the workbook.^30^ This trial found that older hospital patients in the video education group had significantly higher levels of awareness about their risk of falls, as well as better motivation and confidence to engage in falls prevention strategies, compared to those who received the workbook.^30^ These findings from the evaluation of original SRP resources are supported by a review of video-based health education, that demonstrated videos have superior effectiveness in patient uptake and health literacy over other patient educational methods, when presented in an understandable way.^31^

The professional production of the revised video resource was perceived to have better aesthetics and be of higher quality than the original version. The improved reaction to the video contributed to an improved patient viewing experience and consequently improved patient learning. The importance of quality video presentation format has been identified as a key component in the effectiveness of video-based education in a scoping review.^31^ Studies using animated videos or presentations as interventions found consistent improvement in short term patient outcomes such as knowledge and comprehension of information provided by the health care team.^31^

Staff rated the workbook as significantly more engaging and reflective of current clinical practice for fall prevention. This was supported by patient reactions to the workbook as being informative and easy to read. The workbook was intended to be a valuable adjunct to support the video, as it provided an ongoing resource that patients could review in their own time. Staff could also confer with a patient when setting goals, by drawing their attention to relevant workbook content for that individual. The revised program replicated the original SRP workbook by maintaining an adult learning framework and adhering to recommended reading levels for patient education materials.^23, 32^ Readability is recommended to be at a grade six level to allow for low health literacy levels and facilitate patients’ comprehension and subsequent engagement in health decision-making.^23^ However, the revised workbook was smaller, used contemporary pictures, slightly reduced the content and included more overt instruction on goal setting.

Patients participating in the education that used the original SRP resources significantly improved their knowledge of falls prevention strategies, which is consistent with the findings from the original program.^10, 16^ However, patients who received education using the revised SRP showed greater overall improvement. These patients had higher perceived motivation and intention to undertake falls prevention strategies following the revised SRP. Program improvements also led to greater patient awareness of their own personal risk of falls. This is particularly important for two reasons. First, behaviour change frameworks identify that recognising one’s own personal risk of a health problem is a strong motivator for being willing to take action to mitigate risk.^33, 34^ Second, when investigating beliefs and knowledge about hospital falls, older patients, even those at high risk of falls, have been consistently shown to have very low self-perceived risk of falling, as well as low levels of knowledge about falls, and think staff will inform them if they are at risk or required to take action.^8, 35^

### Strengths and limitations

The inclusive participatory approach resulted in the co-production of engaging resources and strongly contributed to the credibility of the SRP revision. The mixed methods design provided triangulation of data and staff and patients had similar positive reactions to the revised SRP, contributing the trustworthiness of the findings. The study used two different cohorts of patients for each phase, rather than have patients do a direct comparison of the two resources. However, this was felt to be necessary and practical by both consumers and staff, as patients needed to be provided with the education in real time as part of the safety procedures of the hospital and hospital staff delivered the education as part of usual care. The study was focused on improving the resources to support better learning by patients and facilitate staff delivery of the SRP. While reactions and learning were improved, health behaviour change theory explains that patients need to set clear behavioural change goals to reduce their falls risk and need high levels of opportunity and motivation to undertake these planned falls prevention strategies.^33, 34^ Therefore, further work to evaluate staff delivery of the revised SRP and the enacting of relevant behaviour change by patients is required. In previous trials, staff delivered the SRP over one to three sessions.^15, 16^ Process evaluations of these trials of the original SRP with this mode of delivery demonstrated that patients who received SRP developed high levels of knowledge, confidence and motivation to engage in personalised falls prevention strategies.^10^ Staff participating in these trials also perceived that the original SRP created a positive culture whereby staff and patients worked together to enact behavioural change around falls prevention and also improved staff knowledge about creating a safe ward environment to engender good support for effective behavioural change.^19^ Hence further trials that identify barriers and enablers to providing the revised SRP as part of hospital falls prevention programs will be valuable to inform implementation of new falls prevention guideline recommendations.^11^

## CONCLUSION

The world falls guidelines recommend that older patients be provided with fall prevention education when in hospital. Education programs should be evidence-based and designed using sound andragogical and behavioural change frameworks. Patient and hospital staff participation contributed to the successful revision of the SRP, an effective individualised fall education program. Both staff and patients showed significant improvements in their reactions to and learnings from the revised program. Patients also demonstrated positive changes in their attitude towards reducing their risk of falling in hospital. Investigating the impact of the revised SRP on older patients’ engagement in fall prevention strategies, and its effectiveness in reducing hospital falls and injuries is warranted.

## Data Availability

All data produced in the present study are available upon reasonable request to the authors.

## Acknowledgement

The authors sincerely thank the staff and patients of Attadale Rehabilitation Hospital for their kind contribution to this research.

